# Fernando de Noronha: how an island controlled the community transmission of COVID-19 in Brazil

**DOI:** 10.1101/2020.10.22.20216010

**Authors:** Mozart Júlio Tabosa Sales, Ligia Regina Franco Sansigolo Kerr, Regina Vianna Brizolara, Ivana Cristina de Holanda Cunha Barreto, Rosa Lívia Freitas de Almeida, Paulo Savio Angeiras de Goes, Luiz Odorico Monteiro de Andrade, Leuridan Cavalcante Torres, Flávia Kelly Alvarenga Pinto, Francisco Marto Leal Pinheiro Júnior, Rebeca Valentim Leite, Aline Priscila Rego de Carvalho, Amanda Carolina Felix Cavalcanti de Abreu, Rebecca Lucena Theophilo, Fernando Rodrigues Magalhães, Susane Lindinalva da Silva, Carl Kendall

## Abstract

**Introduction:** Fernando Noronha (FNA) is a small Brazilian archipelago in the Atlantic, part of the state of Pernambuco that COVID-19 has decimated. Anticipating the worst from the pandemic, Island and state authorities implemented a series of public health actions to contain the epidemic. This paper, reporting the results of the first wave of a cohort study, documents the measures and their effects through a cohort study.

**Methods:** Measures were documented at the time of implementation. A random sample of 904 residents were selected from the health register, interviewed and tested for COVID-19 (RT-PCR and serology). The survey explored socioeconomic variables and adherence to prevention behaviors.

**Results:** Flights were reduced from 38 to once a week, FNA was closed to tourism, schools were closed, and testing and tracing contacts was mandated along with social distancing and use of masks. A household lockdown was briefly imposed for residents. A prevalence of 5.1% was found, and a total of 158 cases of COVID-19 was estimated, although only 28 had been reported in routine surveillance. Half of the population reported food insecurity and applied for government COVID-19 benefits. Adherence to control measures was high, except for intrahousehold mask use with family and friends.

**Conclusion:** Despite high levels of COVID-19 in Pernambuco, continued exposure through the provision of essential services from the mainland, and lack of direction from national authorities, FNA was able to implement a series of prevention measures unique in Brazil that contained the epidemic on the island.

## Introduction

Islands have been a fascination for epidemiologists^1^. The promise of island epidemiology is that transmission can be more closely monitored, and therefore the dynamics of transmission and the effect of interventions can be more effectively explored. This is especially critical in Brazil, until recently, the country with the second largest number of COVID-19 cases in the world^2^. Fernando de Noronha Archipelago (FNA) are 21 islands about 350 kilometers off Brazil’s northeast coast. The official population of the islands is about 3000, all living on the main island that covers 17 km^2^. The total territory of the archipelago is 26 km^2 3^.

FNA is managed by an administrator-general appointed by the Pernambuco state government. Most of FNA was declared a National Park in 1988. In 2001, UNESCO declared FNA a World Heritage Site, boosting tourism, the main economic activity of the island^4^. In 1942, the United States Army Air Force built an airport on the island to support the Allied campaign in Africa. At the end of the war FNA was returned to Brazil. Fernando de Noronha Airport is served by daily flights from Recife and Natal. In January and February 2020, Fernando de Noronha received 38 weekly flights, carrying an average of 452 passengers per day. Between April and June, frequency dropped to one per week, carrying an average of 4 passengers per day ^5^.

Pernambuco is especially hard-hit by COVID-19, registering 132,152 cases (1382.8 cases/100,000 inhabitants) and 7,702 deaths (80.6 deaths/100,000 inhabitants) by September ^6^. FNA initiated prevention activities in the first half of March 2020, before the first official COVID-19 case was reported in the state. These included imposing a lockdown, promoting physical distancing and providing emergency assistance to the neediest families; enhancing testing for Sars-Cov-2, including monitoring of arriving travelers, restricting access to the island and the initiation of the cohort study described here to estimate the incidence and prevalence of Covid-19. In spite of special attention, before the beginning of this study at the end of May, 2020, there were 28 confirmed cases of COVID-19 reported in the archipelago ^7 2020b^. In addition to conventional measures the Government of Pernambuco and local authorities implemented a series of prevention measures unique in Brazil, such as active case detection and contact tracing, and movement regulation for all islanders.

The potential to control the pandemic on this island, and better understand the uptake and effectiveness of control measures motivated our team to focus on Fernando de Noronha. The objective of this article is to discuss these control activities and report the results of the first round of the cohort study.

## Methodology

### Study design and location

We implemented the study in FNA in two ways: 1) documenting pandemic-related events and documents, including epidemiological bulletins; and 2) a prospective cohort study.

First we reviewed data extracted from the following sources: 1) demographic and socioeconomic data from the Brazilian Institute of Geography and Statistics ^8^; 2) state and district decrees and ordinances; 3) number of cases and deaths from COVID-19 reported by the Pernambuco State Health Department; 4) flights and passengers from the National Aviation Agency ^5^; and 5) information provided by local authorities and residents. The data were systematized as per the Center for Disaster Studies, FIOCRUZ ^9^. The second method is a cohort study including questionnaires and biological testing that began on May 22, 2020 and will be repeated at 60, 120, 180 and 360 days from baseline.

### Sample population and size

All individuals residing on the island of any age were eligible to be included in the study. Names and addresses were drawn at random from a current list of all residents. Residents were excluded if not found at home after a second visit and replaced with another randomly selected. The project was described, voluntary participation emphasized, and the Informed Consent Form presented. In the case of minors (<18), parent or guardian provided consent. Ethical review complied with Resolution No. 466/2012 of the National Health Council and was approved by the National Research Commission (Project CONEPE #4.284.892).

While the official population of FNA is 3,061 (IBGE, 2019), local authorities maintain an updated resident health register totaling 4122. Using 4122, a 95% CI, and an acceptable error of 1%, we calculated a sample size of 811. Estimating a loss to follow up of 10%, a final sample of 892 was chosen.

Until April 28, 2020, only 28 confirmed cases of COVID-19 had been notified in FNA ^7^. A national survey estimated that for each COVID-19 case in Brazil, there would be about 15 undiagnosed cases ^10^. However, the situation of the AFN is unique, given exclusion of visitors since March 20. Thus, we estimate that there are about 4 unknown cases for each known case (112 cases), yielding an incidence of 2.7%.

### Data collection

Participants were interviewed with appropriate hygiene measures to record: 1) demographic characteristics; 2) socioeconomic status (SES) and housing conditions, 3) clinical, epidemiological and health services variables; 4) measures adopted to prevent COVID-19; 5) mental health; and 6) food insecurity. To measure SES we used the Brazilian Economic Classification Criteria (BC). BC uses kind and number of possessions, employment status, housing characteristics and education of the household head to characterize SES. Questionnaires were entered using SurveyMonkey® and exported to STATA® v.16 for analysis.

Samples of venous blood were collected for Rapid Serological Test (RST) ^11^ and a nasopharyngeal swab collected for RT-PCR. The RST used the SARS-CoV-2 Antibody Test Kit (IgG / IgM) by Guangzhou Wondfo Biotech Co., Ltd. The viral RNA was extracted using the QuickExtract ™ RNA Extraction Kit (Biosearch. - Ref.: QER090150) according to manufacturer’s instructions. RT-PCR tests were conducted using primers and probes to detect 3 target regions of Coronavirus (N1, N2, N3) and to detect RnaseP. The primer and probe sequences have been validated by the CDC (USA).

### Analysis

A table, listing type and date of intervention and events was prepared. Data from epidemiological bulletins were compared to these measures to explore their effects. For the cohort study, the prevalence of the variables included were calculated using Stata® v.16 with their respective 95% confidence intervals. The findings were compared to national estimates and with experience on other islands of similar size.

## Results

### Control methods for COVID-19

The government of Pernambuco and the FNA administration reorganized the response to COVID-19, opening State and District COVID-19 Offices. The offices promulgated WHO (2020) and PAHO (2020) recommendations concerning social distancing, testing, isolation and other measures. They published state and district epidemiological bulletins about COVID-19 and presented plans and control measures using radio and print. They implemented strict border controls for the island, with drastic reductions in flights. They prohibited entry to the island for almost all civilians, including the re-entry of residents off-island (March 20-May 31).

State Decree (SD) 48,809 / 2020 (03/14/2020) banned events with more than 50 people. SD 48.955 (4/20), quarantined the archipelago, and required authorization for traffic from the district administration. This was restricted to the purchase of food, medicines and health care. Authorization was requested and obtained electronically via cellphone and the administration responded within 24 hours. Evidence of the success of these measures is the registration of 206 permits/day. Using anecdotal information, much of the population remained at home. School activities were canceled on March 18, 2020 (Appendix 1).

The island had no secondary or tertiary care facilities, so primary health care was reinforced, including staff training, measures to reinforce urgent emergency care, and the hiring of additional health professionals. Given the transportation difficulties, a 6-bed Field Hospital was constructed on the island. Surveillance protocols for cases of COVID-19 were established and bulletins and health education materials published. The first case of COVID-19 in FNA was registered on March 27, 2020. Cases detected are placed in quarantine and the evolution of disease monitored daily. The active search of contacts before initiation of the cohort study had been effective in identifying cases. By 04/05/2020, 173 PCR tests were performed in the FNA. By this date, the island had 28 positive cases with three being quarantined and 25 recovered. Another 145 cases were discarded after negative test results (PCR).

In the first week of June, 42 more cases of COVID-19 were identified through the cohort study, totaling 70 cases. During the months of June and July, 17 more cases of COVID-19 were identified by the Health Surveillance System, all of them among essential workers and residents who were returning from the continent, and all were quarantined. By July 30, FNA reached 88 confirmed cases, of which 79 were recovered and 9 were kept in isolation ^7^.

On July 25, entry requirements were modified (ATDENFN, 2020a). As a consequence, from July 26, in order to enter the island, passengers must present a negative RT-PCR test performed no more than 7 days before departure or a positive IgG and negative IgM serological test carried out not exceeding 90 days from the date of mainland departure. In addition, they must comply with the guidelines, such as mandatory use of a mask, maintaining minimum distance, and regular hand washing or use of alcohol gel. Except for those who have presented IgG (positive) and IgM (negative) tests for COVID-19, the quarantine starts from the date of the first test for COVID-19 (RT-PCR) on the continent, until the authorization by the Health Superintendent, which occurs after a second test for COVID-19 (RT-PCR). If this second RT-PCR is negative, the entrant is released from quarantine, if positive, they must remain in quarantine for at least 14 days.

### Baseline cohort results

The final study sample size consisted of 904 residents. Women (52.1%) constituted more than 50% of the sample. The majority of participants were adults (40.5% between 19-39 years and 41.5% between 40-58 years), married or living with a partner (49.4%), less than a third (29.9%) did not complete high school and about 85% belonged to social classes C (44.1%) D or E (39.9%). With respect to work, 82.6% worked before COVID-19 with almost half in a non-formal job. Almost one third of those working lost their jobs due to COVID-19. Approximately one third of the participants (32.1%) lived in a household that participated in a cash transfer programs and more than half of participants reported food insecurity (Table 1). More than half (50.2%) received government COVID-19 benefits.

**Table 1.**
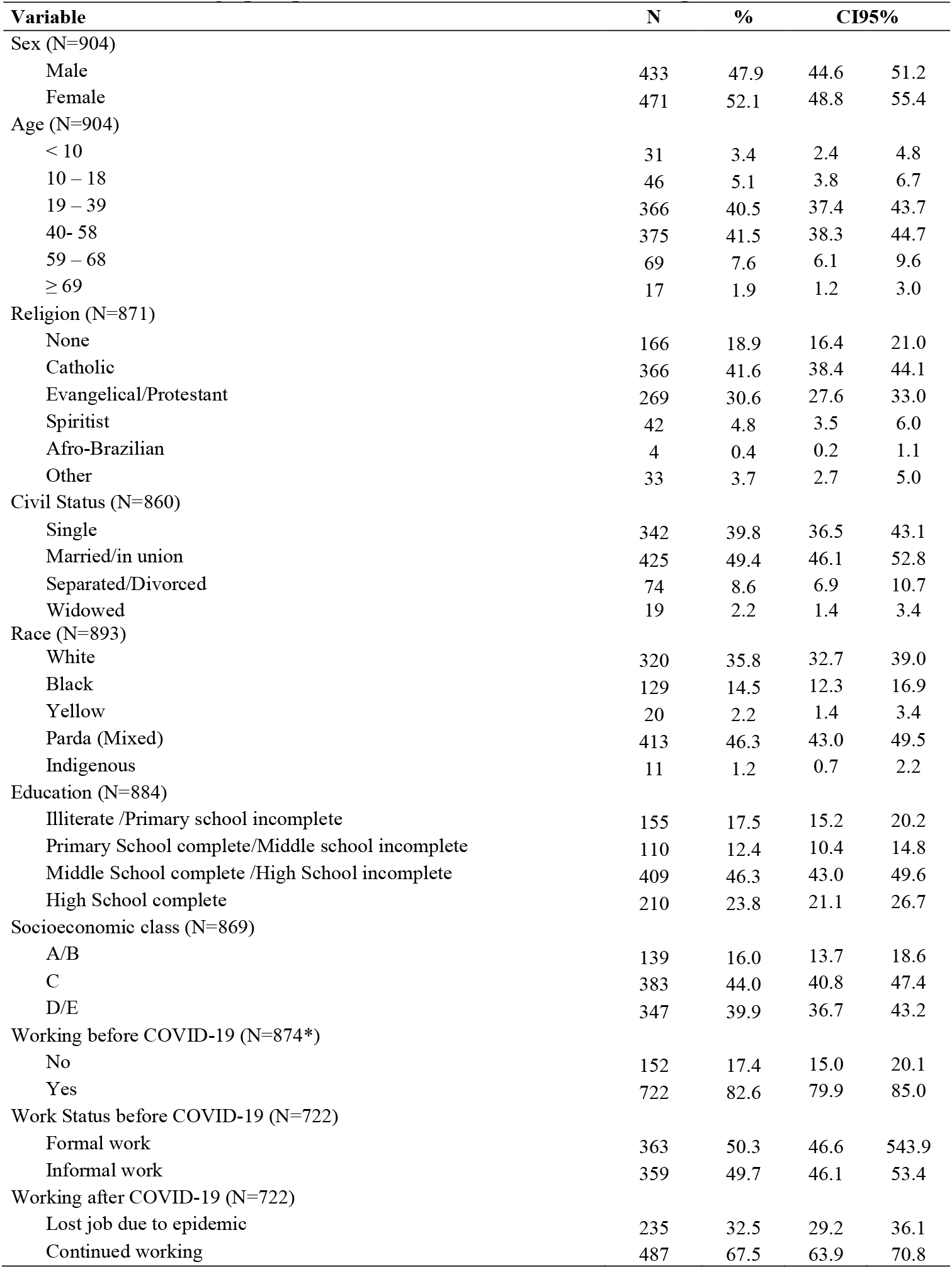
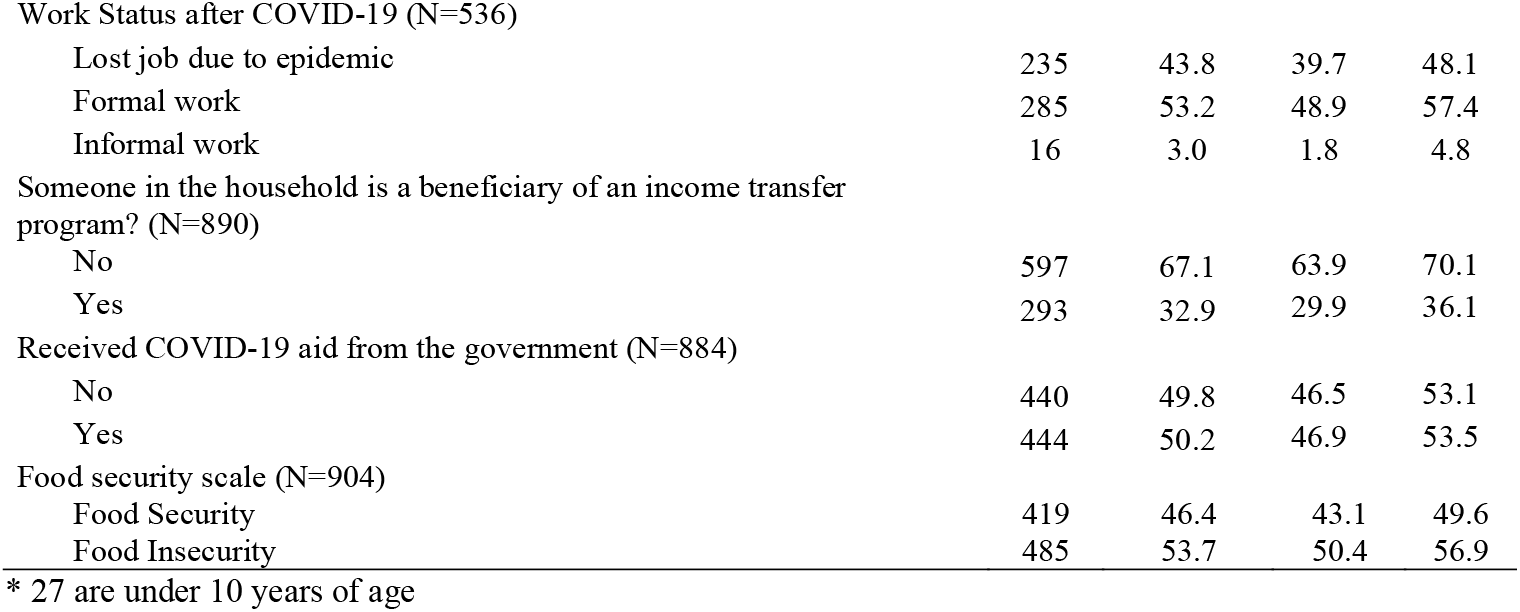
Sociodemographic profile of the Fernando de Noronha sample

While it is difficult to characterize previous and current behavior in a single self-reported questionnaire, answers appeared to demonstrate a selective compliance with regulations: 72.9% reported always washing their hands, 91.7% reported having alcohol in their house, and only 5.3% of respondents reported never leaving home. Reasons for leaving home were essential tasks such as shopping for food (62.6%) and work (39.1%). With respect to masks, 98.7% reported having a mask, and 82.6% reported always using the mask when they left home. However, while about 60% reported that they received visitors (family, friends or delivery personnel) inside their house during the pandemic, fewer always used masks specially when the visitor was family or friend (Table 2). With respect to public behavior, compliance with regulations appears good, although 12.2% of respondents report observing public parties and some crowding in markets and beaches (7.3% and 7% respectively).

**Table 2.**
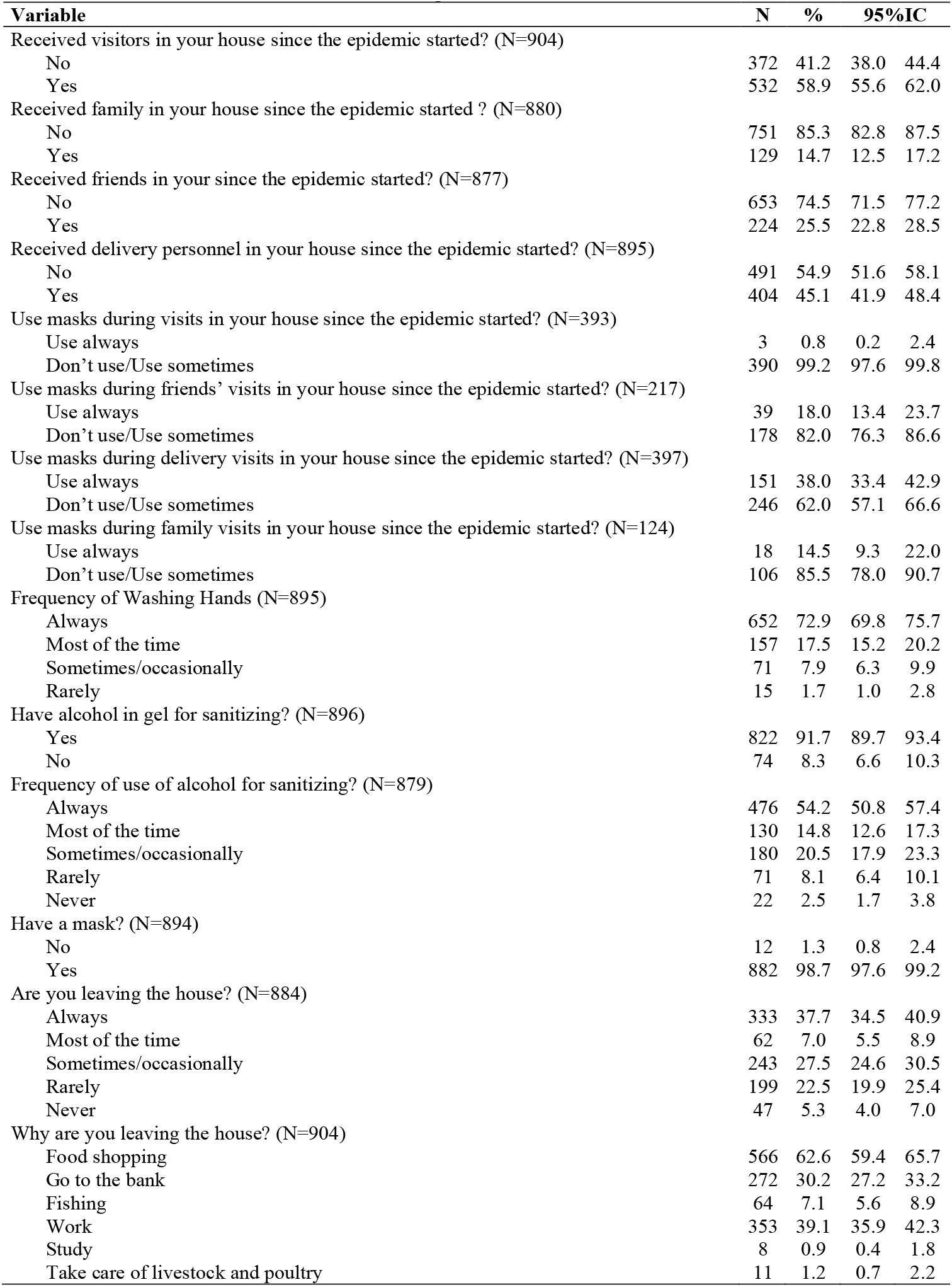
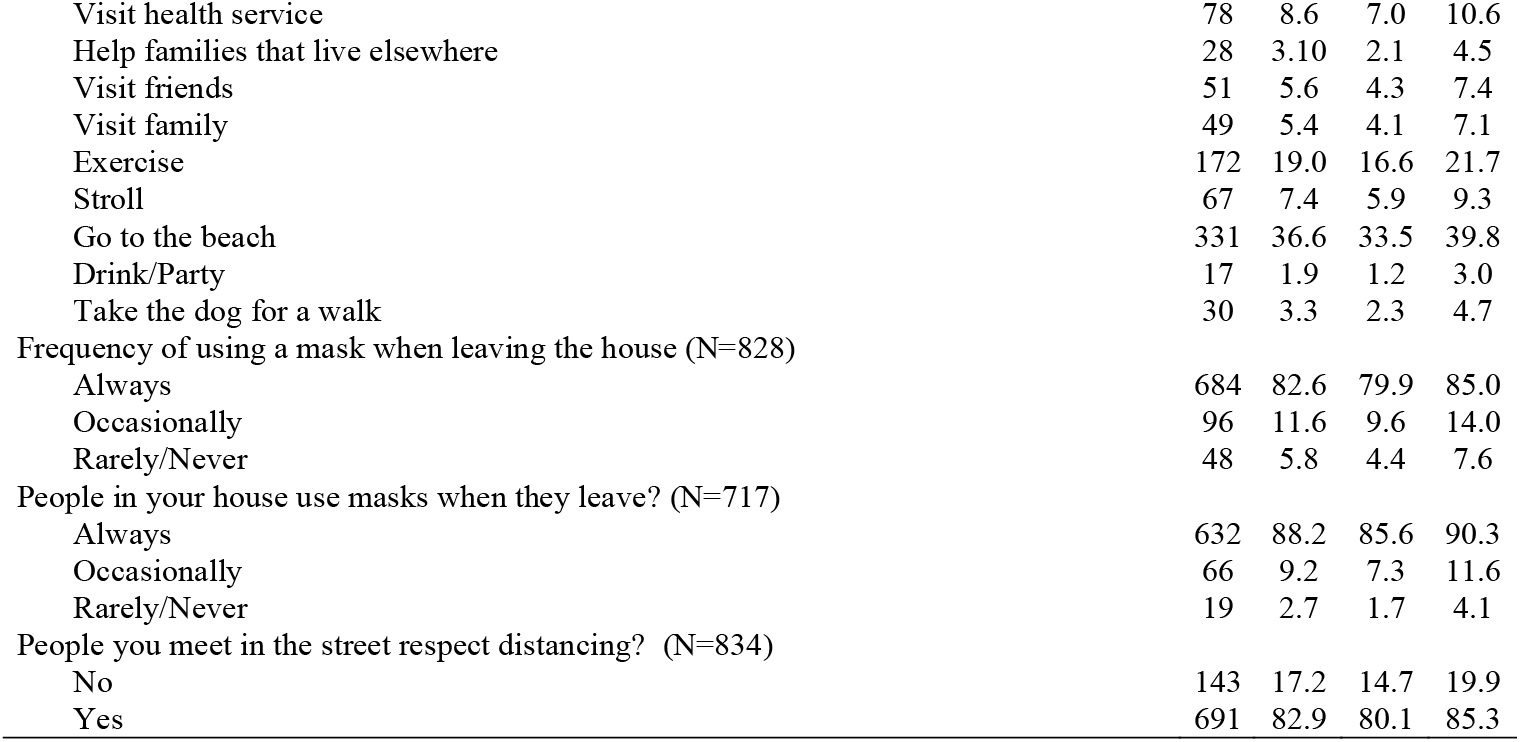
Pandemic related behaviors among FNA residents.

The survey reports a combined COVID-19 prevalence rate of COVID-19 (TSR and/or RT-PCR) of 5.1% (95% CI: 3.8-6.7), and an incidence of 1.0% (95% CI: 0.5 - 2.0) measured by RT-PCR (Table 3). The TSR positive rate was 4.3% (95% CI: 3.2-5.9). Among those 46 positive participants, only 5 had been previously identified. Thirty-nine participants tested positive for TSR; 9 tested positive for RT-PCR, with 7 only positive for RT-PCR, 34 positive only for TSR and 2 were positive for both TSR and RT-PCR. Participants who were RT-PCR positive underwent quarantine, were retested and contacts investigated. No new cases were identified through contact tracing. There was no significant difference in prevalence or incidence between genders. By age group, children under 10 showed the highest percentage of previous infection: 8.6% (95% CI: 2.7-23.8) were positive for RRT; 5.9% (95% CI: 1.4-21.1) for RT-PCR and 14.3% (95% CI: 6.0–30.4) for both. There was no RT-PCR positives among participants aged 10 to 19 years and those 60 or older.

**Table 3.** Serological and RT-PCR results of participants in the first wave of the cohort in Fernando de Noronha Island by sex and age group, May 2020.

| L1 | % | IC95% |  |
| --- | --- | --- | --- |
| TSR<sup>1</sup> ou RT-PCR (N=904) |  |  |  |
| Positive (n=46) | 5.1 | 3.8 | 6.7 |
| Negative (n=858) | 94.9 | 93.3 | 96.2 |
| TSR<sup>1</sup> (N=904) |  |  |  |
| Positive (n=39) | 4.3 | 3.2 | 5.9 |
| Negative (n=865) | 95.7 | 94.1 | 96.8 |
| RT-PCR (N=879) |  |  |  |
| Positive (n=9) | 1.0 | 0.5 | 2.0 |
| Negative (n=870) | 99.0 | 98.0 | 99.5 |

<sup>1</sup> Rapid Serological Test

Half of the patients positive for COVID-19 (50.0%; 95% CI: 35.8 - 64.2) and 17.7% (95% CI: 15.2 - 20.4) of the negatives reported that they became ill. All symptoms investigated were more prevalent in the participants who tested positive for COVID-19. More than 40% (95% CI: 28.6 - 57.1) of positive cases lived with someone positive for COVID-19 in the same household, while only 7.1% (5.5 - 9.0) of negative cases reported the same (Data not shown).

## Discussion

This study demonstrates that the measures taken in FNA successfully interrupted community transmission of COVID-19 on the island about three months after the first detected cases. This was the case even though the epidemic was raging on the mainland, especially in Pernambuco. While Sars-Cov-2 has spread rapidly around the world, infecting more than 30 million people and causing the death of almost 1 million (Sept 19, 2020), some islands have been “spared” the pandemic’s advances. Islands attached to the United States in the Caribbean, specifically Puerto Rico and the US Virgin Islands, have not been successful in controlling the epidemic ^12^ or controlling air access to the island. On the other hand, 12 nations in the world, 10 in the Pacific, report they have not had a single case of COVID-19 ^13^. Part of this is due to geography and the abrupt interruption of flights around the world, providing some isolation, as was the case with the FNA.

However, if we take into account the official population of the island ^3^ and the 5.1% prevalence reported in our survey of the island, we would estimate 158 cases of COVID-19 (95% CI ranging from 118 to 208 cases) actually occurred on the island, although only 28 had been reported, that is, 5.6 times more cases than those identified in routine health surveillance. Comparing the prevalence of COVID-19 in Fernando de Noronha with the results of the national survey of 133 cities (EPICOVID19-BR) released on May 20, 2020, the FNA would have had the ninth highest prevalence rate ^10,14^.

The FNA employed several exceptional public health measures that were not used elsewhere in Brazil, such as: 1) contact tracing and testing for all positives identified on the island; 2) for travelers, testing (RT-PCR) within seven days before departing for FNA; 3) RT-PCR testing on arrival to the island and quarantine until authorization to leave (if negative) or extended for 14 days if positive; 4) use of a wrist band to identify positives that could only be removed by the island’s health surveillance system; 5) and travel restrictions on the island for residents, with the use of a cell-phone app to request authorization to leave the house, similar to the card used in China ^15^. The measures adopted in the archipelago were a good example of a more restrictive social distancing policy at the beginning of the country’s pandemic ^16^.

Despite the measures discussed in the previous paragraph, transmission was relatively high, probably due to one or a combination of three factors. The first considers adherence to prevention measures which are reported at levels higher than for any of the nine states in Northeast. ^17^ However, although always use of mask outside the home was high, use inside the home was not, especially for friends and extra-domestic family members. A second potential transmission route is extra-island transport, such as food and other products arriving by air or ship. While there are protocols for handling cargos, such as fuel, packages and crew are not always subjected to them. Transporting food and other cargo is a necessary feature of a small island, especially one dedicated to tourism. Handling, delivery, unpacking, and preparation for use present many potential points of transmission. A third factor is that the island requires a number of off-island specialized staff, and has a military presence. Specialized staff maintain the electrical grid and serve as fire fighters, and the military maintains a continuous presence on the island. This requires a greater effort on the part of epidemiological surveillance to monitor this floating population who are frequently moving between the island and the continent. It also requires close and consistent communication with the organizations responsible, in order to achieve compliance with COVID-19 prevention protocols.

We found that unemployment and informal work increased on the island after COVID-19. An important proportion of those who worked lost their jobs and others had to resort to informal work, without rights or benefits. When the COVID-19 pandemic started, Brazil already had 41% of its workers in informal positions and since the pandemic unemployment rates have increased in 12 Brazilian states, half in the Northeast, including Pernambuco ^18^. The situation of the FNA is peculiar and aggravated by the strong dependence on tourism as an economic activity. It is estimated that 100 million jobs were lost as a result of the COVID-19 pandemic around the world ^19^ and the impact has been greater on islands. For one example, the British Virgin Islands report that 92% of its GDP is linked to tourism ^19^. Even with a rebound expected in the future, the ability of the FNA economy to recover may be affected by many factors: low levels of tourism, availability of flights, vaccine efficacy and availability, declining or rising case numbers and other determinants of visa restrictions for tourists.

The situation of joblessness and work in the informal sector are associated with rising food insecurity, with more than half of the residents reporting food insecurity in our study. The results of the Family Budget Survey conducted by IBGE in the 2027-2018 biennium, when the government that assumed the presidency implemented profound reductions in expenditures, shows that, at that time, more than a third of Brazilian households presented some degree of food insecurity, the highest index registered since 2004 when the survey was conducted for the first time ^20^. Of the 68.9 million households in Brazil, 36.7% had some degree of food insecurity, affecting 84.9 million people. In addition, the worst situation was recorded in the North and Northeast regions, where more than half of the households reported food insecurity. This correlates with our results on the island, where only 46.4% reported food security. The pandemic could only make the situation worse, especially in the absence of a national government that acted strongly in meeting these socioeconomic needs ^21,22^. Other studies in Brazil have shown a positive correlation between low family income per capita and unemployment with the incidence of COVID-19, which reinforces the importance of cash transfer programs ^23^. The aid proposed by the federal government, at the beginning of the pandemic, was around US $37 per month, that when pressured by the Brazilian Congress, was raised to US $111, which is still considered too little to support families.

An important editorial highlighted damaging reactions and inadequate responses from leaders of countries, as in Brazil, who denied the seriousness of the disease and subsequent deaths.^24^ Studies have evaluated the uninformative role of many opinion leaders in Brazil about COVID-19 on social media^25^, demonstrating the negative impact on social distancing and other recommended measures.^26^ This illustrates one effect of the fragmentation of policy and program among the federal government, state and local levels and helps explain the difficulties faced during the pandemic in Brazil^17^.

## Conclusion

Part of FNA’s success in controlling COVID-19 could be attributed to the isolation provided by its geography, but we believe that it was largely due to a series of measures taken by local authorities that led to the control of community transmission. The FNA is a successful example of disease control in Northeastern Brazil - contrary to what has occurred in other states in the country. Among the strategies that deserve to be highlighted are the tracking and early identification of cases, extensive testing, isolation of positive and suspected cases, and the travel protocol for landing on the island, practices not systematically implemented in any part of the country. These are activities that can be continued on the island during the next phases of the epidemic.

However, sustaining this example of success will be a challenge after the opening of the island to unrestricted travel and tourism. The population of the island, due to its’ dependence on tourism and the serious situation of unemployment, underemployment and food insecurity, continues to pressure local authorities to reopen tourist activities on the island.

The future of controlling the disease will depend on how well current practices can be ramped up to control new cases during the reopening of the island as well as how well the global, regional and national economic recovery can respond to the worsening social and economic situation on the island. The cohort study presented in this article will continue to monitor the island response and provide information for local decision-makers.

## Data Availability

This is a cohort study. When the study and analysis are completed, the dataset will be made available.

## Acknowledgements

We thank the Pernambuco State Health Department; Fernando de Noronha District Authority, and Itaú’s “Todos pela Saúde” initiative for funding. We also thank Dr. Socorro Gros, representative of the Pan American Health Organization / World Health Organization in Brazil; Dr. André Longo, State Secretary of Health of Pernambuco; Pedro Ribeiro Barbosa, CEO of the Institute of Molecular Biology of Paraná; Guilherme Cavalcanti da Rocha Leitão, Administrator of the Fernando de Noronha District Authority; Tereza Campos and Afra Suassuna, respectively General Superintendent and Research Superintendent of the Instituto de Medicina Integral Fernando Figueira; and to the health professionals of Fernando de Noronha, who were instrumental in carrying out the fieldwork in this research.

## Contributions

Conception (C); Data collection (DC); Analyses (A); Writing paper (WP)

**C, A, WP**: Mozart Júlio Tabosa Sales; Ligia Regina Franco Sansigolo Kerr; Regina Vianna Brizolara; Ivana Cristina de Holanda Cunha Barreto; Paulo Savio Angeiras de Goes; Luiz Odorico Monteiro de Andrade; Leuridan Cavalcante Torres; Flávia Kelly Alvarenga Pinto **DC, WP**: Rebeca Valentim Leite; Aline Priscila Rego de Carvalho; Amanda Carolina Felix Cavalcanti de Abreu; Rebecca Lucena Theophilo; Fernando Rodrigues Magalhães; Susane Lindinalva da Silva

**A, WP:** Rosa Lívia Freitas de Almeida; Francisco Marto Leal Pinheiro Júnior

## Appendix

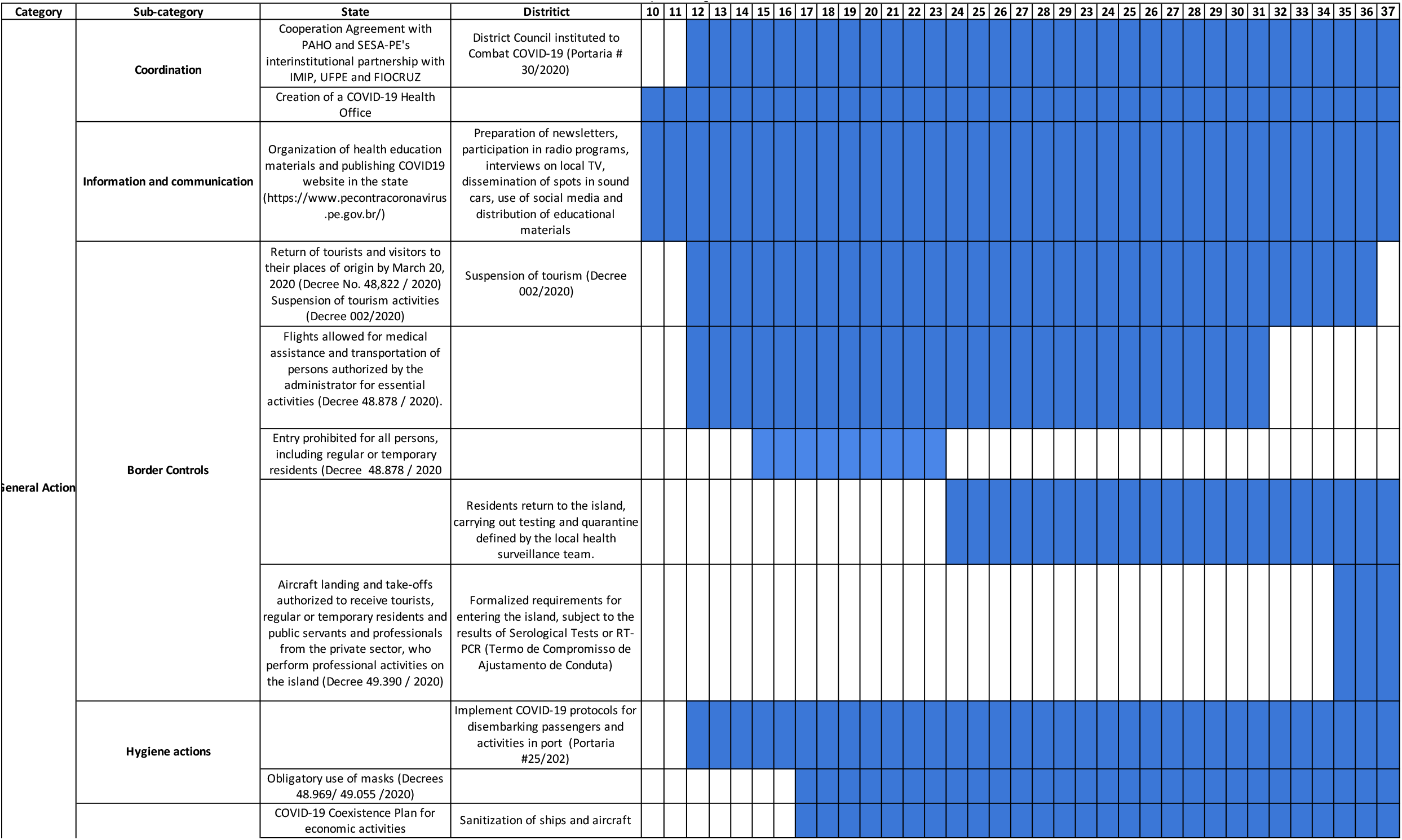
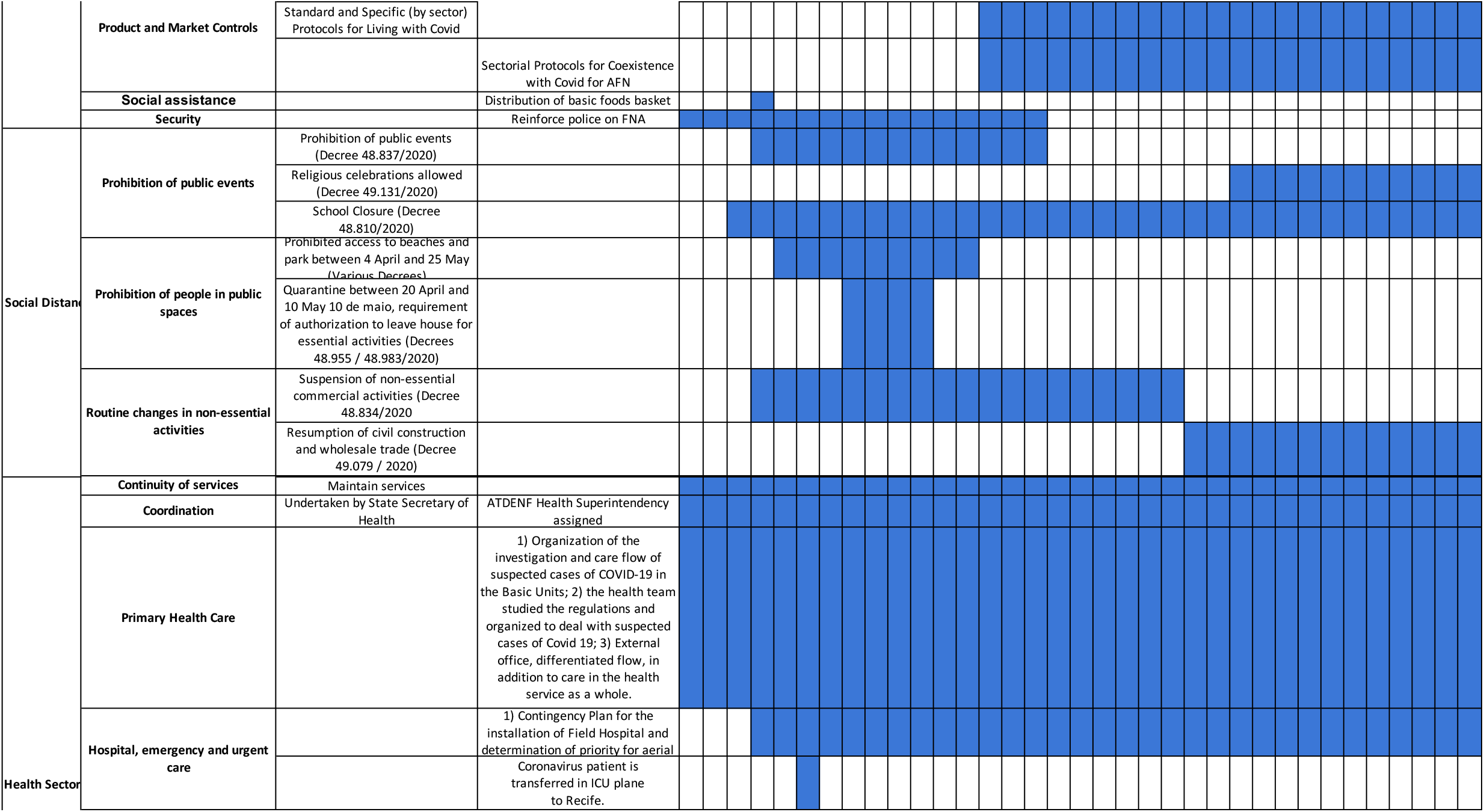
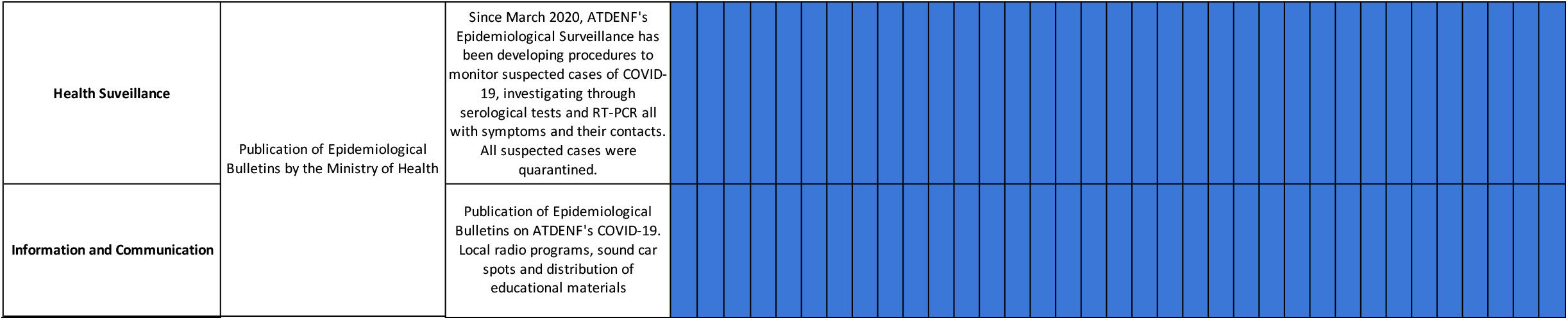

## Notes

### Competing Interest Statement

The authors have declared no competing interest.

### Funding Statement

Funding for data collection was provided by Pernambuco State Health Department; Fernando de Noronha District Authority, and Itau’s Todos pela Saude initiative. No other funds were received.

### Author Declarations

The project was approved by the IRB of the "Instituto de Medicina Integral Professor Fernando Figueira - IMIP" in Pernambuco with approval number 4.112.781. The IRB of IMIP is approved by the Brazilian National Ethical Committee (CONEP).

